# Advance of Novel Coronavirus Registration Clinical Trial

**DOI:** 10.1101/2020.03.16.20034934

**Authors:** Gao Song, Meng Qun Cheng, Xian Wen Wei

**Author notes:** These authors contributed equally to this work (Meng Qun Cheng, Xian Wen Wei). Corresponding author: Gao Song. Department of Pharmacy,The Puer People’s Hospital of Yunna City, 44 Zhenxing Rd., Puer 665000, China.

## Abstract

**Background:** To analyze the characteristics and heterogeneity of clinical trials of Novel Coronavirus(COVID-19) registered in the China Clinical Trial Registry (ChiCTR), and provide data bases and information references for clinical treatment

**Methods:** Statistics of COVID-19 clinical trials registered with ChiCTR as of February 24, 2020 were collected. Descriptive analysis of registration characteristics. The chi-square test is used to compare statistical differences between different study types, intervention methods, study stage, and Primary sponsor.

**Results:** 232 COVID-19 studies registered at the ChiCTR were collected. The overall number of COVID-19 registrations was increased. Hubei Province, China has the largest number of registrations. There were significant differences between the number of participants(P=0.000), study duration(P=0.008), study assignment(P=0.000), and blind method(P=0.000) for different study types. Significant differences could be seen in the dimensions of multicenter study(P=0.022), of participants numbe(P=0.000), study duration(P=0.000) and study assignment(P=0.001) for the four intervention methods. There were significant differences in study assignment(P=0.043) between the early and late studies. CMT drugs with high research frequency are chloroquine, lopinavir / ritonavir, and I-IFN; BI was Cell therapy, plasma therapy, Thymosin, and M/P-AB.

**Conclusions:** Different study design characteristics have led to significant differences in some aspects of the COVID-19 clinical trial. Timely summary analysis can provide more treatment options and evidence for clinical practice.

## Background

Since the 1970s, more than 30 new types of infectious diseases, such as Ebola virus disease, human infection with highly pathogenic avian influenza, and Zika virus disease, have appeared in China, seriously threatening people’s health and public health security [1,2]. The Novel coronavirus (COVID-19) is a new highly pathogenic coronavirus that broke out in Wuhan, China in December 2019. It can cause severe pneumonia and is listed as Public Health Emergency of International Concern (PHEIC) [3].

As of February 25, 2020, China has accumulated more than 70,000 confirmed cases of Novel coronavirus pneumonia(NCP), which have been reported internationally in 24 countries and 5 continents [4]. The timely conduct of clinical drug research and the promotion of clinical trial information sharing are critical to epidemic prevention and control.In May 2006, the World Health Organization (WHO) officially launched the International Clinical Trials Registry Platform (ICTRP). In July 2007, The ChiCTR was officially certified by the ICTRP as a first-level registrar, becoming the fourth primary registrar after the Australian/New Zealand Center, the American Center, and the British Center [5]. According to the Declaration of Helsinki and the regulations of National Medical Products Administration (NMPA), all medical research conducted in China must be registered in ChiCTR [6]. As of February 24, 2020, the ChiCTR Center has registered more than 200 COVID-19 clinical studies. It is of great significance to discuss the research status and development situation in this field in time, which is effective for conducting clinical research and accelerating the transformation of results.

Therefore, this study collates and analyzes the registration of COVID-19 clinical studies to provide an information-based data foundation and information reference for curbing the spread of COVID-19 epidemic.

## Methods

### Data collection

Check the “COVID-19 Clinical Research Index” issued by the ChiCTR Center, search the ChiCTR database with the keywords “Novel coronavirus” or “2019-nCoV” and count the registration information as of February 24, 2020. Extract the following information: (1) Registration status (date of registration, date of completion, region of registration, etc.). (2) Source of funds. (3) Recruitment status. (4) Ethical approval. (5) Data Management Committee. (6) Research types. (7) Research design. (8) Research stage. (9) Study time limit. (10) Number of participants. (11) Intervention methods. (12) Setting Blinding.Two reviewers (Gao Song and Mengqun Cheng) were transferred to SPSS and checked to determine the final dataset. There was no disagreement between the two reviewers about the dataset with 232 studies.

### Definitions

Basic research characteristics: research type, intervention method, funding source and research stage. It is defined as: (1) Type of research: intervention, observation, prevention, diagnostic test, prognosis research. (2) Intervention methods: Chemical treatment (CMT); Bio logics and immunoregulatory drugs (BI, such as cell therapy, antibodies and glucocorticoid); Traditional Chinese Medicine Treatment (TCM, such as Chinese herbal medicine, proprietary Chinese medicines); Behavioral intervention (BEI, such as cognitive, attitude, and behavioral interventions, exercise and psychological therapy, etc.); no (missing information or not applicable [NA]). (3) funding source: industry (company, company), public (hospital, university, government), self-funding. (4) Research stage: early stages (stage 0, I, 1/2, 2), late stage (stage 3 or 4), and non-applicable stage. (5) Ethical approval: yes, no. (6) Data Management Committee: yes, no.

Study design characteristics: multicenter study, number of participants, study duration, study assignment, Blinding, and key inclusion/exclusion criteria. It is defined as: (1) Number of study locations: Single study location or Multiple study locations; (2) Number of participants: <100, 100 to 300, 301 to 1000,> 1000. (3) Study duration(month): <1, 1 to 3, 4 to 12,> 12. (4) Blind method: open, single blind, double blind, unspecified. (5) Study assignment: randomized, non-randomized, factorial grouping, continuous grouping, and others. (6) key inclusion/exclusion criteria: yes.

### Data analysis

Descriptive analysis of registration time trends and geographical distribution, the registration volume reflects the overall trend of COVID-19 clinical research. Descriptive statistics were calculated to investigate the percentage distribution of all of the items studied.The frequency calculation of multi-center research institutions is based on the statistics of the responsible units. The study duration was calculated by ChiCTR’s “Study execute time”.Chi-square tests at the 95% significance level were then performed to study possible differences in design characteristics for research type, intervention method, funding source and research stage. In the chisquare test, we only statistically analyze the main features, excluding some feature analyses. Additionally, as 100.0% of the 232 studies had key inclusion/exclusion criteria, these items were ruled out in the chisquare tests. Statistical analysis was performed using SPSS22.0 software. P<0.05 was considered statistically significant.

## Results

### COVID-19 clinical study registration

A total of 232 COVID-19 clinical studies were registered with ChiCTR. Overall, the number of registrations has increased steadily (Figure 1).The number of registrations before February 1 was small, and the number of registrations quickly increased to 8 on February 2, reaching a peak on February 14(n=18).On January 23, 2020, China’s ChiCTR Center registered the first COVID-19 clinical study, and the intervention method was CMT treatment (ChiCTR2000029308).The first TCM clinical study was registered on January 27 (ChiCTR2000029381). The first BI study was registered on February 1 (ChiCTR2000029431). The BEI study was first registered on February 2.

**Figure1.**
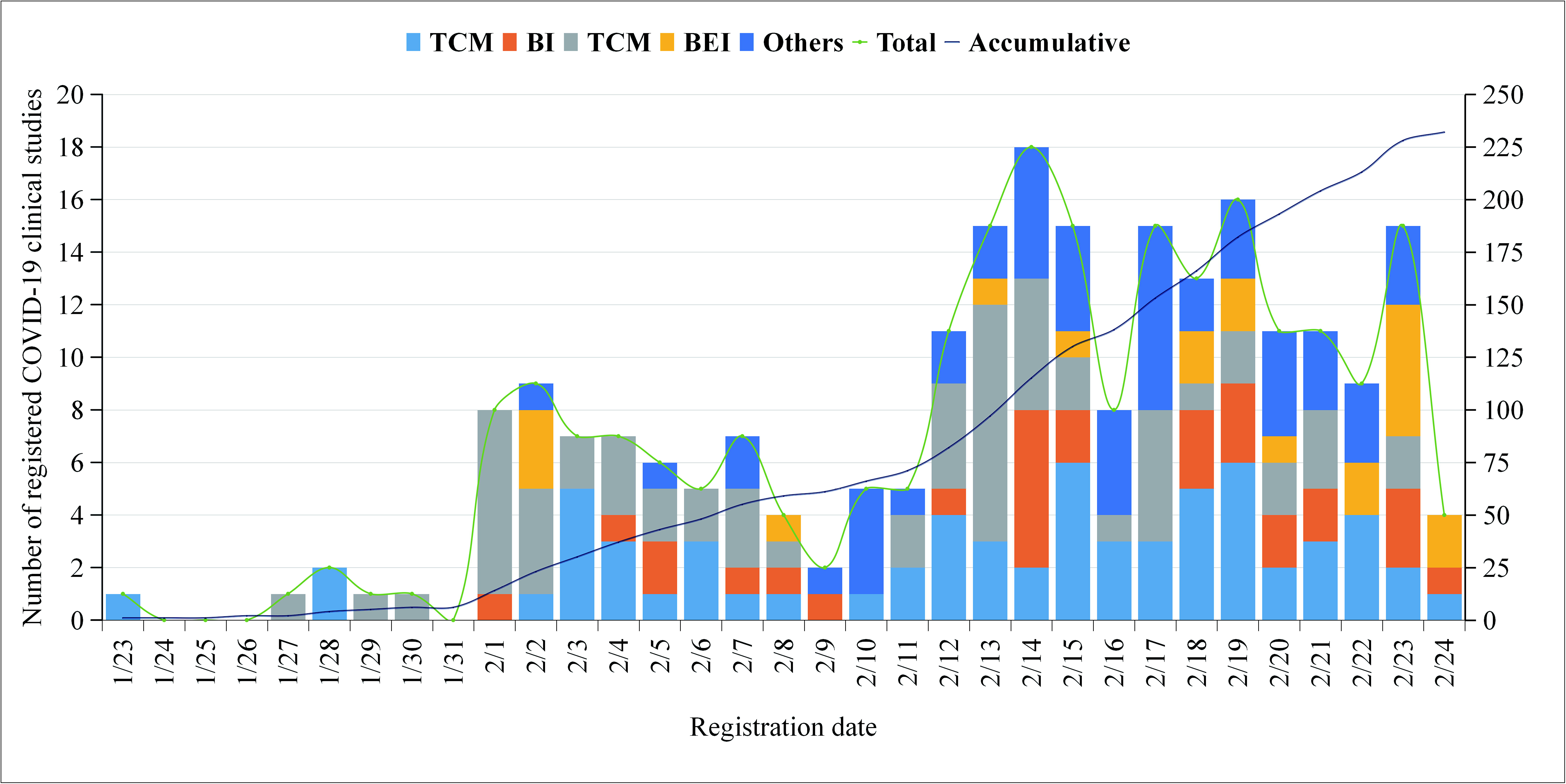
Trend of registration time for COVID-19 clinical studies.

As of the statistical date, a total of 18 provinces and 4 municipalities nationwide participated in the COVID-19 clinical study (Figure 2).The national registration volume ranked first in Hubei Province (n=69, 29.74%), followed by Guangdong Province (n=29, 12.50%), and Zhejiang Province and Sichuan Province tied for third place (n=23, 9.91%).Tongji Hospital, Tongji Medical College, Huazhong University of Science and Technology is the most registered hospital(n=16).

**Figure2.**
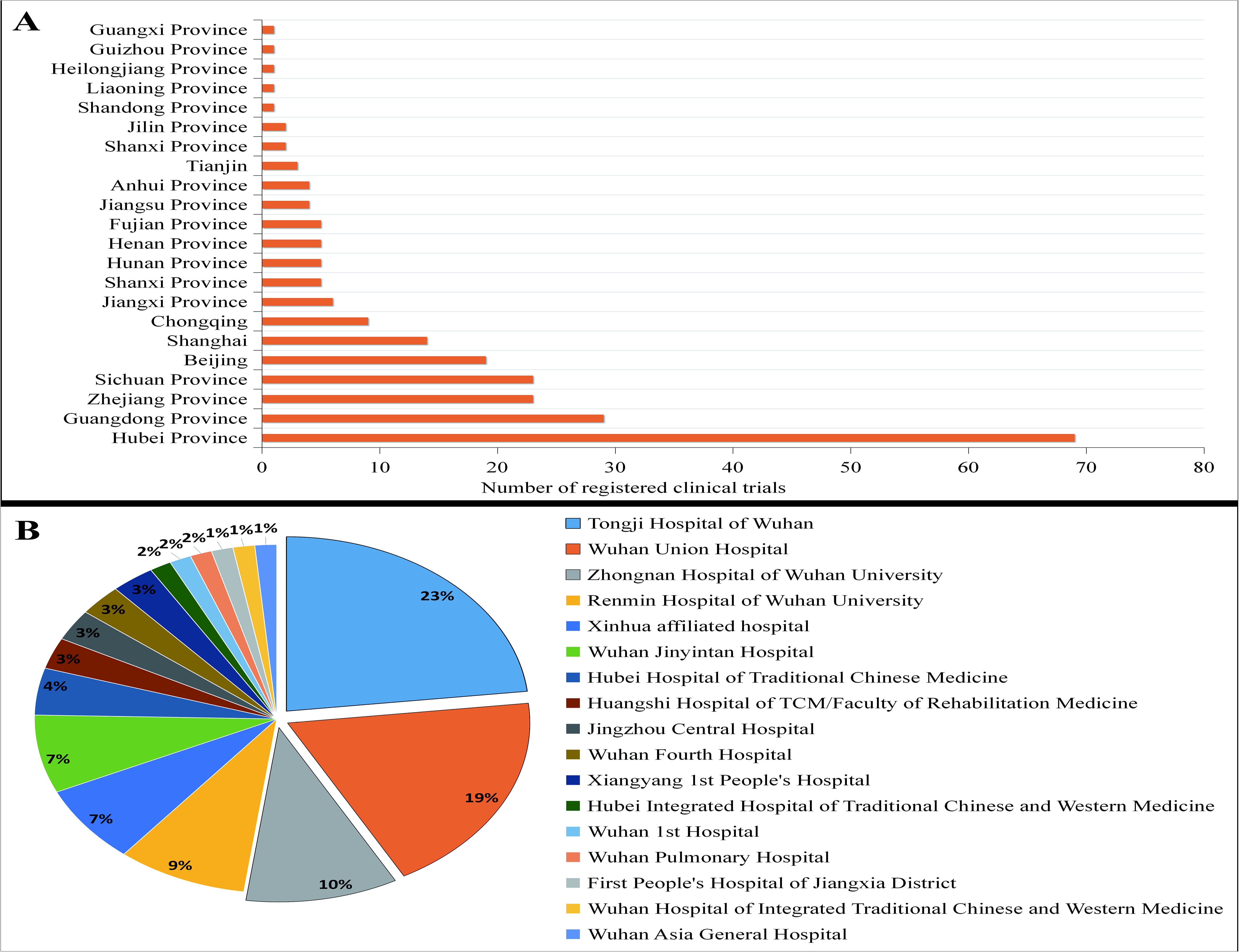
Regional distribution of registration institutions for COVID-19 clinical studies.(A)Provinces and municipalities registered for COVID-19 clinical research in China;(B)Registered Unit of Hubei Province.

### Heterogeneity analysis of study design features

As shown in Table 1, 67.67% of the studies were intervention studies and 25.00% were observational studies. 28.02% of the studies were treated with CMT, 12.93% were treated with BI, 28.02% were treated with TCM, and 8.62% were treated with BEI.The early stage accounted for 41.81% of all studies, and 20.69% of the studies were late stages. 53.88% of research is mainly funded by the public, and 57.76% of research is being recruited. However, 22.84% of the studies did not receive ethical approval, and 47.41% were not confirmed by the data management committee.

**Table 1.** Basic characteristics of COVID-19 registered clinical trials

|  |  | N=232 |
| --- | --- | --- |
| Type of Study |  |  |
|  | Interventional | 157(67.67%) |
|  | Observational | 58(25.00%) |
|  | Diagnostic experiment | 12(5.17%) |
|  | Prognose study | 3(1.29%) |
|  | Preventive research | 2(0.86%) |
| Intervention model |  |  |
|  | CMT | 65(28.02%) |
|  | BI | 30(12.93%) |
|  | TCM | 65(28.02%) |
|  | BEI | 20(8.62%) |
|  | N/A | 52(22.41%) |
| Stage of Study |  |  |
|  | Early stage ( 0 , I , II ) | 97(41.81%) |
|  | Late stage ( III , IV ) | 48(20.69%) |
|  | Not applicable stage | 87(37.50%) |
| Primary sponsor |  |  |
|  | Industry | 10(4.31%) |
|  | Public | 125(53.88%) |
|  | Self-paying | 97(41.81%) |
| Recruiting status |  |  |
|  | Actively recruiting | 134(57.76%) |
|  | Not yet open for recruitment | 98(42.24%) |
| Ethical approval |  |  |
|  | Yes | 179(77.16%) |
|  | No | 53(22.84%) |
| Data Management Committee |  |  |
|  | Yes | 122(52.59%) |
|  | No | 110(47.41%) |

As shown in Table 2, only 6.47% of the studies were multicenter studies. 44.83% of the study participants were between 100 to 300, and only 3.45% of the studies were >1000. 43.53% of the studies were completed within 4-12 months, and only 2.16% were completed within 1 month. 75.0% of the studies used random assignments, but 70.69% of the studies lacked information on blinding.

**Table 2.**
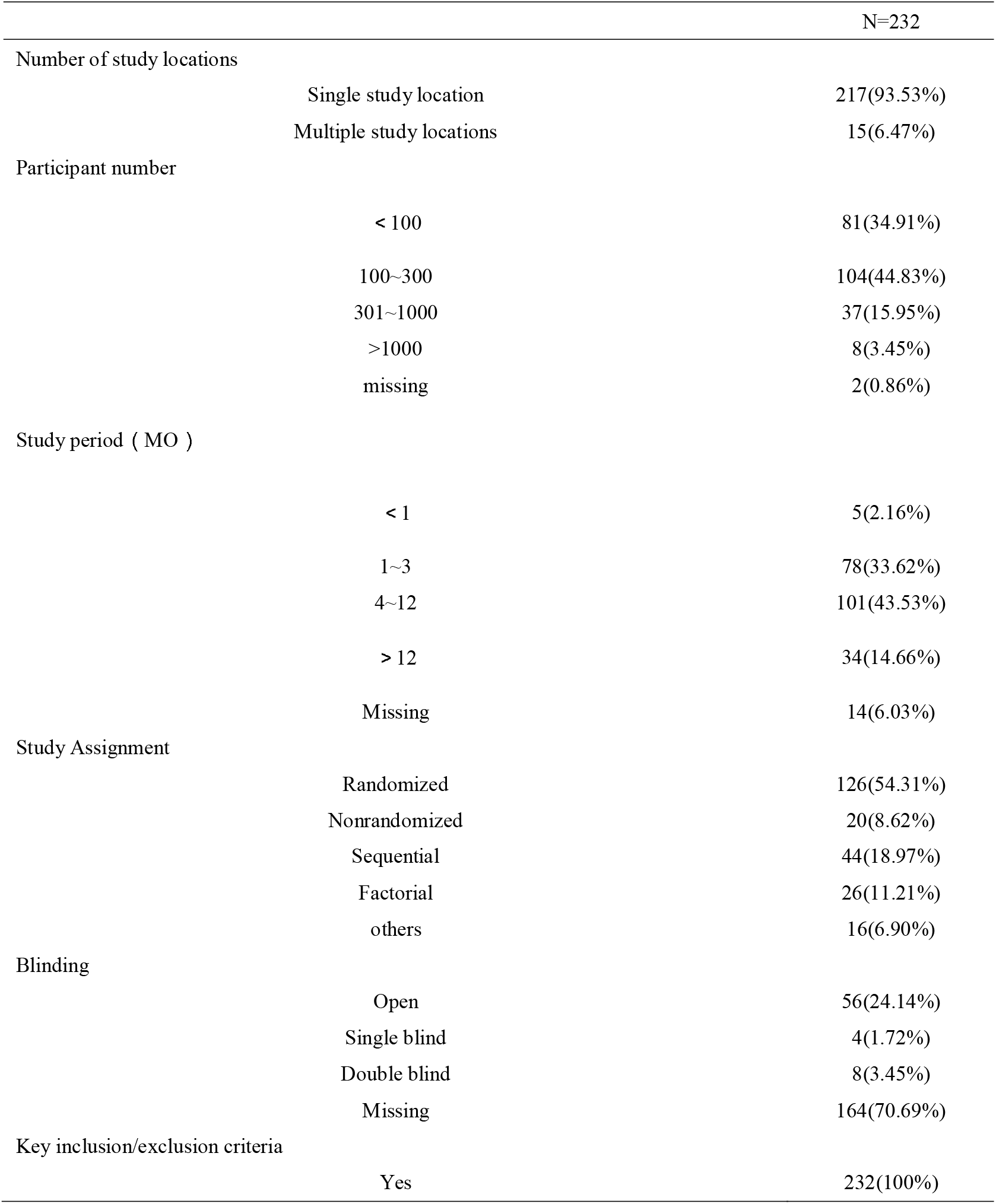
Design features of COVID-19 registered clinical trial

As shown in Table 3,the chisquare tests revealed significant differences in the design characteristics, depending on the Type of Study, for the items of participants number(*P*=0.000), study duration (*P*=0.008), study assignment (*P*=0.000) and blind method (*P*=0.000). The number of participants in 45.86% interventional and 43.10% observational studies was concentrated in the 100-300 range.46.50% interventional and 37.93% observational studies were completed within 4-12 months. 78.98% of the interventional studies were randomly assigned to the group, and 58.62% of the observations were continuous. Only 7.01% of interventional studies were blinded.

**Table 3.**
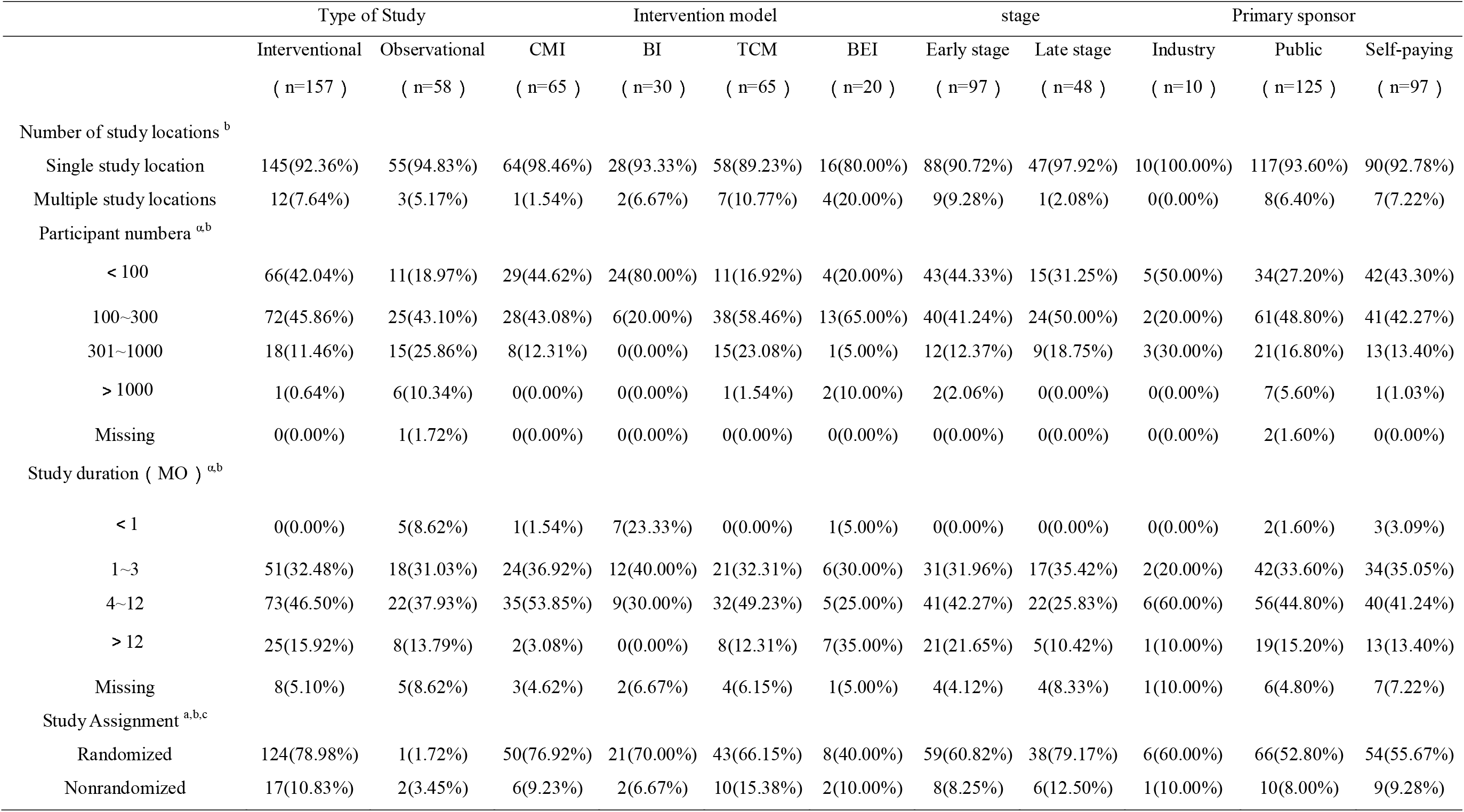
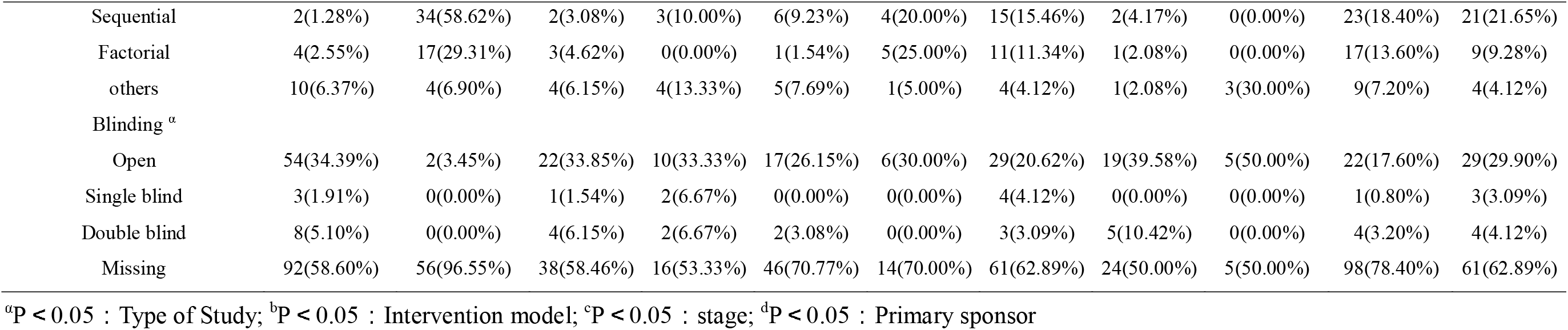
Design characteristic differences among Basic characteristics of COVID-19 registered clinical trials

Significant differences could be seen in the dimensions of multicenter study (*P*=0.022), of participants numbe (*P*=0.000), study duration(*P*=0.000) and study assignment(*P*=0.001) for the four intervention methods.Only 1.54% of CMT studies are multi-center studies. Conversely, 20.00% of BEI studies are multi-center studies, accounting for the largest proportion.The number of participants mainly accounted for: 44.62% of CMT subjects were less than 100, 80.00% of BI subjects were less than 100, and 58.46% of TCM and 65.00% of BEI subjects were between 100 to 300. The study period mainly focused on: 53.85% of the CMT studies were completed within 4 to 12 months, while 40.00% of the BIs were within 1 to 3, 49.23% of the TCMs were within 4 to 12, and 35.00% of the BEI studies were longer than 12 month.For study assignments, 76.92% of CMT, 70.00% of BI, 66.15% of TCM, and 40.00% of BEI studies were randomized. Only 7.69% of CMT, 13.34% of BI, and 3.08% of TCM were blinded in the study.

There were significant differences in the dimensions of study assignments (*P*=0.043) in the early and late study stage.60.82% of the early stages and 26.21% of the late stages were randomly assigned to the group. However, there were no significant differences in the main research dimensions of different funding sources (*P*>0.05).

### Analysis of the characteristics of CMT and BI research

All drugs involved in CMT and BI treatment research are summarized, and the frequency of research is recorded (the same drug is only recorded once in the same clinical trial).68.97% of interventions were drug treatment (n=160). In the CMT study, the top three drugs studied by frequency: Chloroquine (n=16, 16.84%), Lopinavir/ritonavir (n=15, 15.79%), and I-IFN (n=11, 11.58%).

In the BI study, the top three drugs in the study frequency were: cell therapy (n=15, 48.39%), plasma therapy (n=5, 16.13%), Thymosin (n=2, 6.45%) and M/P-AB (n=2, 6.45%). Among them, 32.26% of Cell therapy is mesenchymal stem cells, 9.68% of Cell therapy is Cord Blood Mononuclear Cell, 3.23% of Cell therapy is Menstrual Blood-derived Stem Cells, and 3.23% of Cell therapy is Natural Killer Cell.Figure 3, 4 and Table 4 for a summary of the high research frequency drugs for CMT and BI.

**Figure3.**
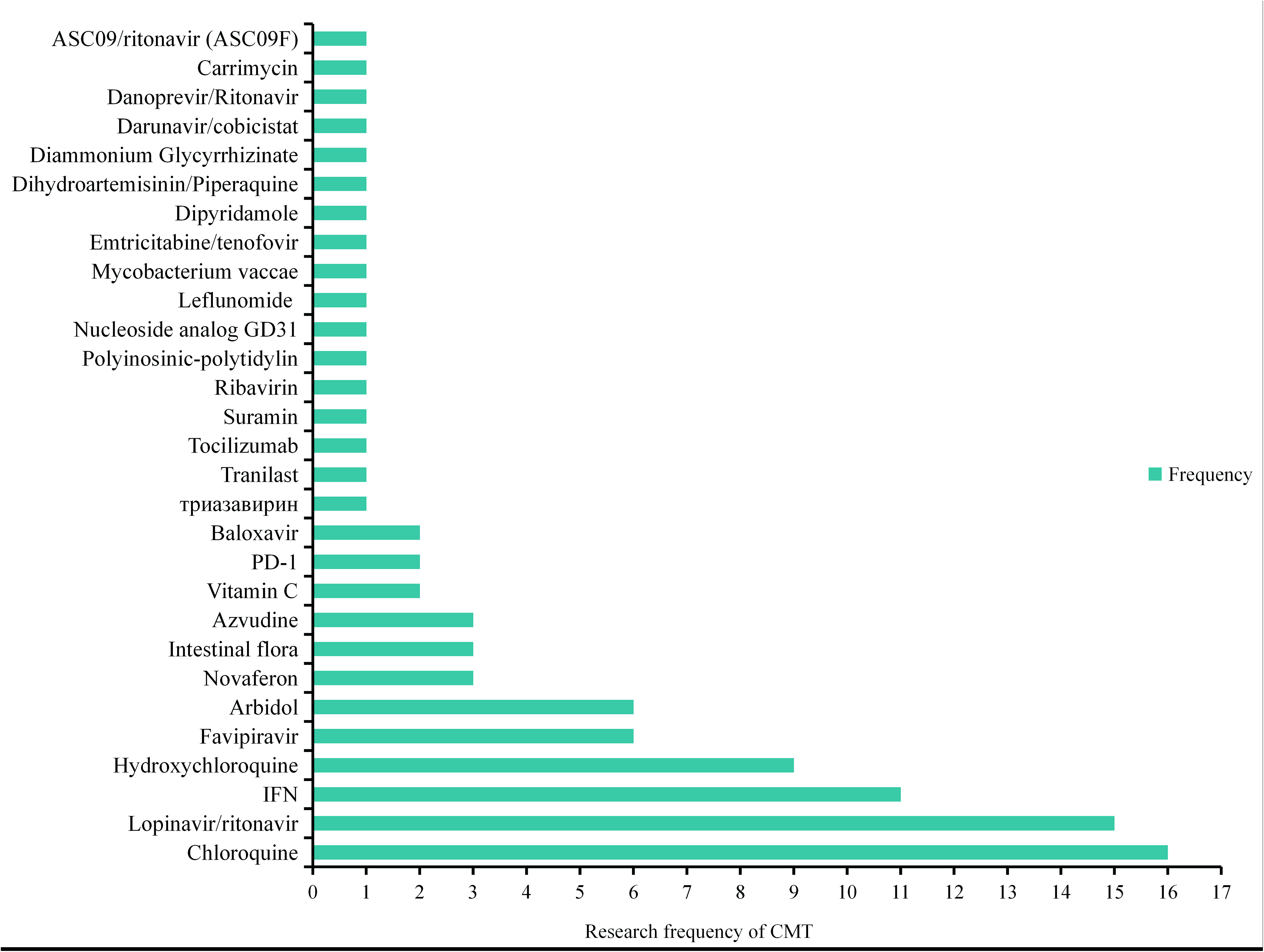
Statistics chart of research frequency of CMT therapeutic drugs.

**Figure4.**
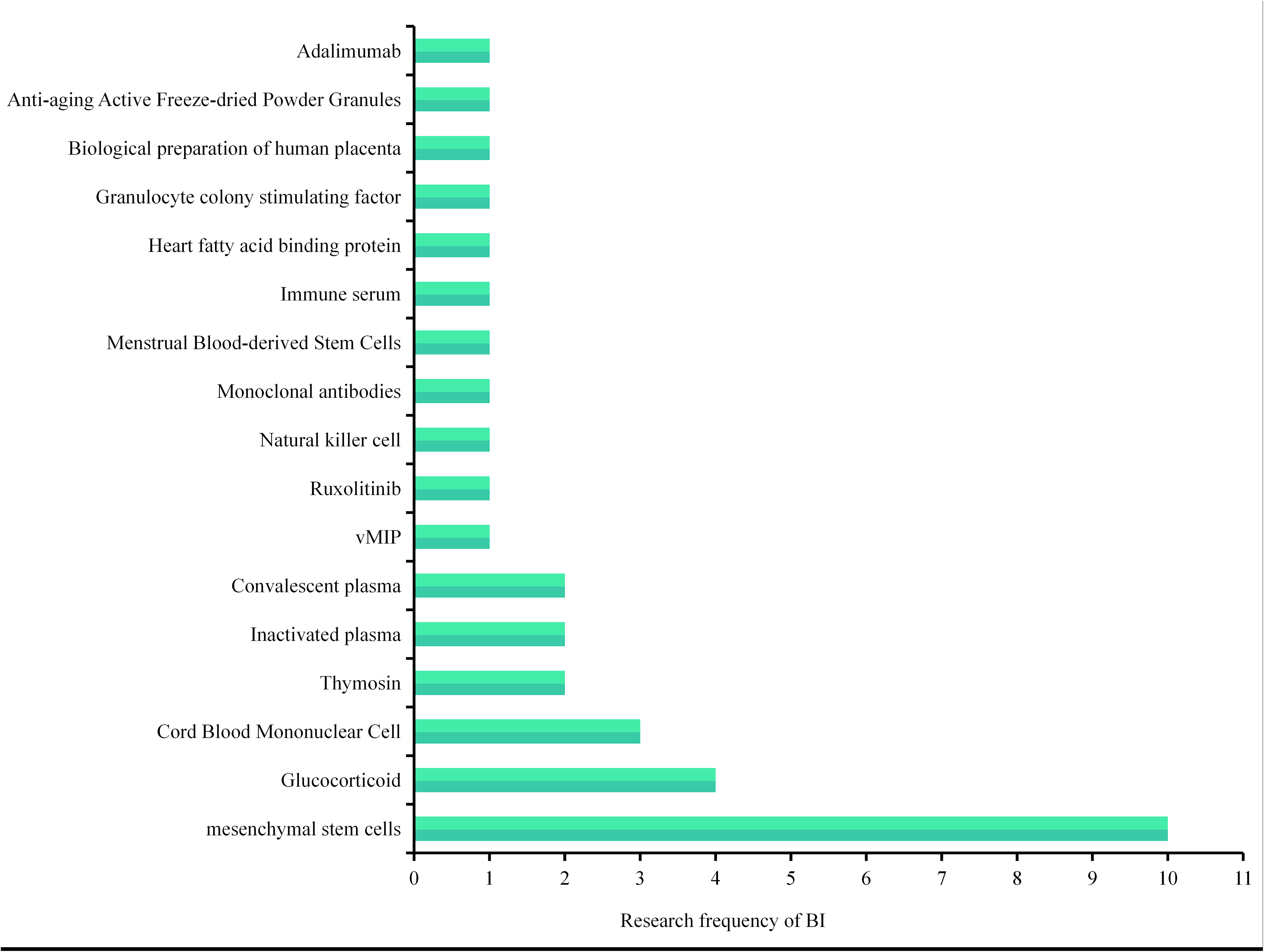
Statistics chart of research frequency of BI therapeutic drugs

**Table 4.**
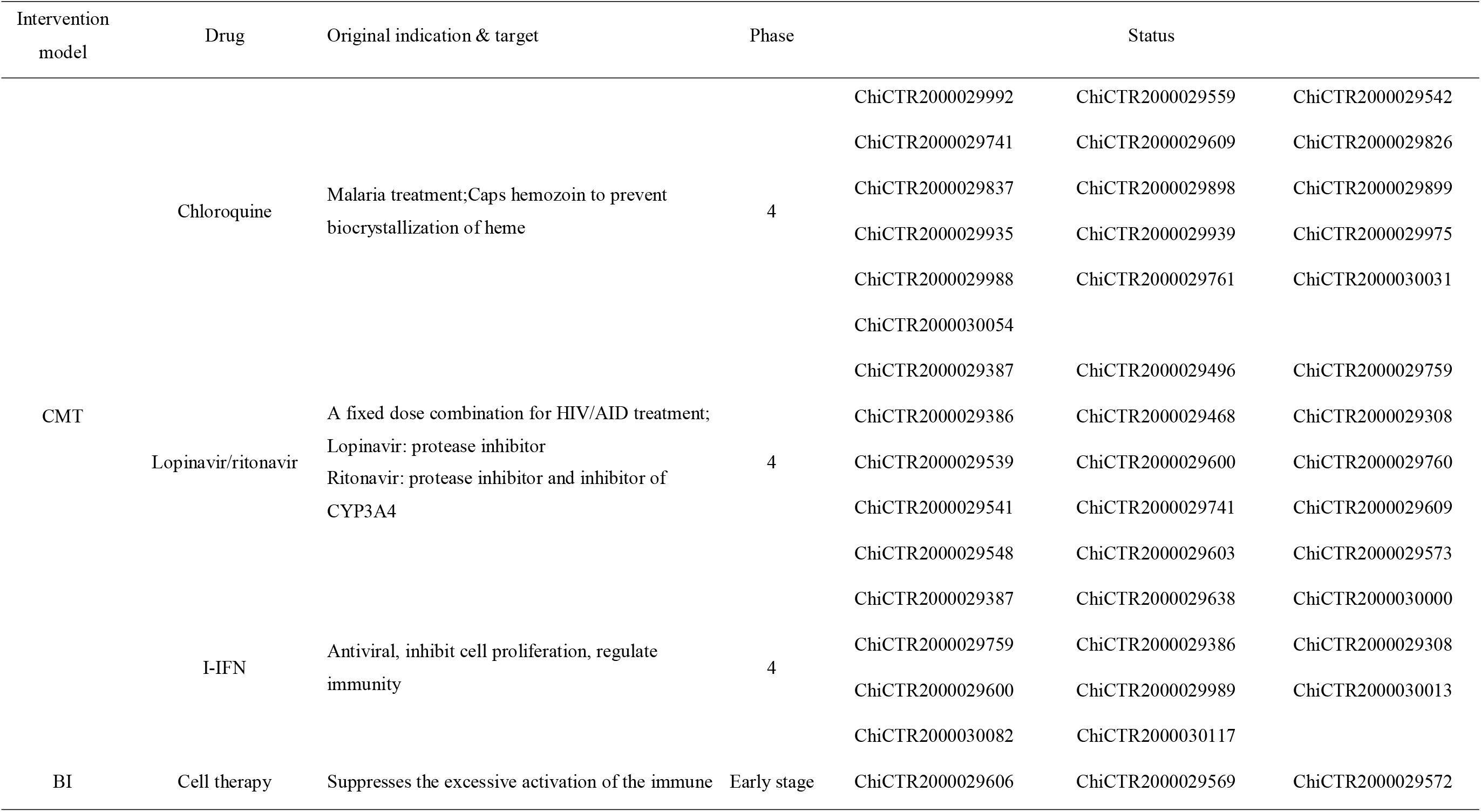
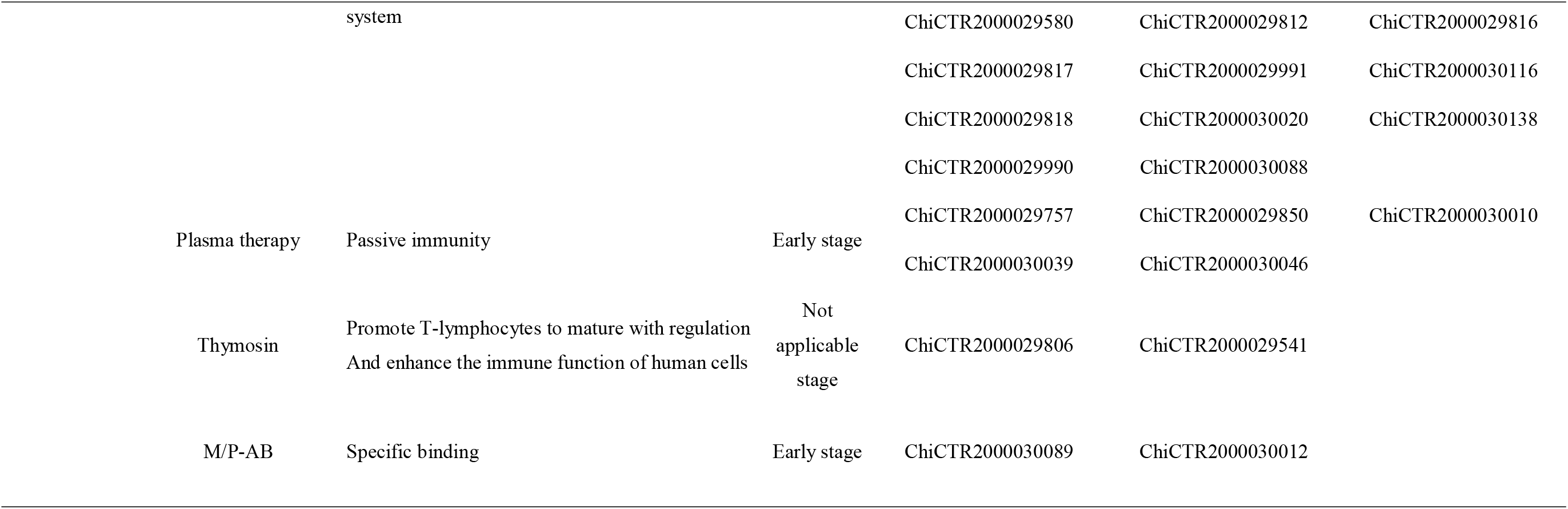
CMT and BI studies in the clinical registry for the treatment of COVID-19 infection

## Discussion

Achieving information sharing in clinical trials is the key to accelerating the transformation of clinical resources and promoting scientific breakthroughs[7]. In the face of a public safety crisis such as an outbreak, the timely sharing of information is even more important, and it can provide real-time guidance for preventing and controlling the epidemic, assessing development trends and the effects of intervention measures[8].COVID-19’s “war epidemic” and “epidemic prevention” strategies are closely related to the important measure of improving the transparency of clinical trials, including sharing of the subject’s original data and standardizing trial design, registration, and implementation [9].

At present, China’s clinical trial registration policy is to implement registration[10], which requires registration in accordance with the ChiCTR registration guidelines. The ChiCTR registration guidelines divide research types into 7 categories, such as interventional studies, preventive studies, diagnostic tests, and observational studies, etc.[11]. However, it is ambiguous that the registration guidelines classify studies by their nature and purpose. According to international standards, it may be more clear to classify intervention and observation [12].

In order to regulate registration, more details should be provided on the randomization procedure and blind method. We found that most interventional studies use randomized allocation, but there are low blinding rates and lack of information. The possible reason is that it is difficult to implement blind method in epidemic situation, but it may also be incomplete information. More than half of the studies did not indicate blind method information. Missing information means that no information about the size of the study is available, which adversely affects the transparency of registration[13]. The results showed that more than half of the studies were not confirmed by the data management committee. Low blinding rate and lack of information may be related to it’s. The detection or guidance of the data management committee should be further strengthened to improve the quality of research design[14].

It is worth noting that 22.84% of the studies failed ethical verification.All human-oriented clinical trials must in any case follow the “Helsinki Declaration”, which has become a consensus of the medical community, and follow the basic ethical principle of “doctors should provide medical care in the best interests of patients” to ensure the subjects[15,16].The ChiCTR Center has issued a warning and requested the supplementary information of the failed units to be reviewed.Medical ethics review should strictly abide by the review work content of the “Ethical Review Methods for Biomedical Research Involving Humans”[17], and here are two suggestions: (1) undertake experimental projects that match actual capabilities; (2) design of experimental schemes An appropriate research method must be selected based on the nature of the problem to be solved.

As of February 24, 2020, the overall number of COVID-19 registered clinical studies is on the rise. The possible reason is that China has already initiated a first-level response to major public health emergencies, and conducting relevant clinical trials is critical to epidemic prevention and control. In addition, in order to cooperate with the epidemic prevention and control work, many national drug clinical trial institutions(GCP) in China have established detailed work guidelines and guidelines to effectively speed up clinical trial review. From the statistical analysis of China’s geographical distribution, it is found that the number of registered provinces in Hubei Province ranks first, and the number of registered provinces close to it is higher than that of other provinces. The possible reason is that the COVID-19 epidemic was mainly concentrated in Wuhan, Hubei Province, China. There was no large-scale epidemic outbreak in remote provinces, and there were relatively few NCP patients.

We found significant differences in the dimension of study design characteristics for different study types.The results showed that compared to observational studies, most interventional studies had fewer than 100 participants. However, 8.62% of observational study designs were completed within 1 month. Such designs were not reflected in interventional studies, and most of the interventional studies were completed in 4-12 months. Similarly, compared to observational studies, most interventional studies use randomized allocation. These results imply that different interventions have an impact on the design of clinical trials.

The design of COVID-19 clinical trials should give priority to “timeliness”. The trial sample size needs to consider the balance between clinical and statistical significance, and its estimated volume reflects the reliability and repeatability of the research results[18].As public health emergencies do not allow more time for exploratory clinical research and there is no historical data, the sample size estimation of such trials may encounter unprecedented problems.Some scholars have proposed[19], whether it is possible to reduce the required sample size by “reducing the power” or “increasing the type I error rate” to complete the clinical trial as soon as possible?But such an approach requires a variety of trade-offs between regulators, sponsors, and researchers.

In addition, the study period is also “time-effective”. If a clinical study is long, it may not make much sense in terms of public health emergencies to respond to the outbreak. From the statistical data, it can be seen that the time limit for completing most of the clinical studies of COVID-19 is concentrated within 4-12 months, and very few studies are completed within 1 month. The clinical research to deal with the epidemic is to race against the virus and the epidemic. This time limit seems too long. Is it possible to conduct mid-term analysis or other adaptive, new design methods? Clinical trial statistics need to be continuously developed based on actual needs.

In addition to the prevention and control of the epidemic, vaccines and drugs are the two major weapons to overcome the epidemic.The previous SARS-CoV epidemic, the 2009 influenza epidemic, and the Ebola epidemic in West Africa were basically approved for marketing at the end of the epidemic, with a lag[20,21].Similarly, the development of specific drugs is also very difficult.The “old medicine” is a new way to increase the indications, and conducting clinical research on anti-COVID-19 may be an important means to respond to the current epidemic.

The “New Coronary Virus Pneumonia Diagnosis and Treatment Scheme (Trial Version 6)” (hereinafter referred to as the “Scheme”) states that there are currently no effective drugs to treat NCP patients.

Drugs such as lopinavir / ritonavir, interferon, abidol, and chloroquine phosphate can be tried, but their effectiveness needs to be confirmed in clinical studies. In the registration trial, the main treatment was drug therapy, accounting for 68.97%. Among them, there are more CMT and TCM studies, and relatively few BI studies. The most frequently studied CMT is chloroquine phosphate.Chloroquine phosphate is an antimalarial drug marketed for many years, and it has been reported in the literature[22] that it can interfere with the glycosylation of the ACE2 receptor, thereby exerting antiviral effects.In vitro experiments show that chloroquine phosphate has the activity of inhibiting COVID-19, and its half effective concentration (EC50) is 1.13 μ/moL[23].As of February 24, 16 clinical studies of chloroquine phosphate-based NCP have registered clinical trials with ChiCTR. Chloroquine phosphate may have clinical therapeutic value.

Mesenchymal stem cells are the most frequently studied in immunotherapy (n=10). Immune abnormalities are the main reason for the progression of patients with severe new-type coronavirus pneumonia. MSC can regulate both the innate immune system and the acquired immune system, and has become the most promising treatment for immune disorders[24,25].MSC can regulate inflammation through a series of mechanisms, including inhibiting T cell hyperproliferation, inducing CD4^+^CD25^+^FoxP3^+^regulatory T cell (Tregs) subsets, and also inhibiting B cell hyperproliferation, differentiation, and immunoglobulin production[26]. At the same time, it can also regulate the secretion of major inflammatory factors and anti-inflammatory factors[27].Immunotherapy of MSC may have clinical therapeutic value.

The “Scheme” emphasizes the important role of TCM. The treatment of the “pathogenic factor” the mechanism by which TCM treats the cause of COVID-19[28]. In addition, its multi-target regulation of the body’s immune system has a therapeutic effect on “cytokine storm” [29]. Currently, relevant clinical studies have confirmed the effectiveness of TCM[30], but further validation by large-scale RCT is needed. Chinese scholars[31] have counted the research frequency of TCM, this article does not count.

BEI research also plays an important role. The rapid implementation of BEI intervention strategies in the early stages of the epidemic can effectively control the spread of the epidemic[32-35]. The main strategies are: (1) Early effective risk communication, timely understanding of the relevant knowledge, knowledge, attitudes and behaviors of the public, and health education. (2) Restrict the travel of people in key areas and mobilize the whole society.

This study has some limitations. The data for this study were derived only from ChiCTR. There may be other databases containing COVID-19 clinical studies. The time limit for research registration is short, and relevant registration information has not been filled in, and it cannot truly reflect its true quantity and level. However, the innovation of this article is to summarize and analyze the registration of ChiCTR’s COVID-19 clinical research in a timely manner, reflecting the research status and development trend of clinical trials in this field, which is “time-effective”.

## Conclusion

There were problems of unclear classification of research types and irregular registration behavior. Also, within the studies researched, heterogeneity exists for various dimensions. Different research types, intervention methods, and research stages lead to significant differences in some dimensions of the COVID-19 study. Finally, statistical high-frequency research drugs can provide more treatment options and evidence-based evidence for the clinical practice.

## Data Availability

All data are from China's CHICTR, the data is true. http://www.chictr.org.cn/index.aspx

**Abbreviations**
COVID-19: Corona virus disease 2019;
ChiCTR: China Clinical Trial Registry;
PHEIC: Public Health Emergency of International Concern;
NCP: Novel coronavirus pneumonia;
ICTRP: international Clinical Trials Registry Platform;
CMT: Chemical treatment;
BI: Biologics and immunoregulatory drugs;
TCM: Traditional Chinese Medicine Treatment;
GCP: National Drug Clinical Trial Institutions;
I-IFN: I-Interferon;
PD-1: CD279;
M/P-AB: Monoclonal/Polyclonal Antibodies;
vMIP: Virus macrophage inflammatory protein;
hFABP: Heart fatty acid binding protein;
G-CSF: granulocyte colony stimulating factor;
BPHP: biological preparation of human placenta;
FDPG: Anti-aging Active Freeze-dried Powder Granules.

## Acknowledgements

Not applicable.

## Authors’ contributions

XWW and MQC contributed to research design. GS and MQC search and extract data from the database. GS helps with data analysis. GS and MQC drafted the manuscript. GS and MQC revised the manuscript. The final draft read and endorsed by all authors

## Availability of data and materials

Not applicable.

## Ethics approval and consent to participate

Not applicable.

## Consent for publication

Not applicable.

## Author details

1 Department of Pharmacy,The Puer People’s Hospital of Yunna City, 44 Zhenxing Rd., Puer 665000, China.

2 Department of Reproductive Medicine,The Puer People’s Hospital of Yunna City, Puer 665000, China.

3 Department of Neurology,The Puer People’s Hospital of Yunna City, Puer 665000, China;

